# Bayesian modeling for the detection of adverse events underreporting in clinical trials

**DOI:** 10.1101/2020.12.18.20245068

**Authors:** Yves Barmaz, Timothé Ménard

## Abstract

**Introduction:** Safety underreporting is a recurrent issue in clinical trials that can impact patient safety and data integrity. Clinical Quality Assurance (QA) practices used to detect underreporting rely on on-site audits, however adverse events underreporting remains a recurrent issue. In a recent project, we developed a predictive model that enables oversight of Adverse Event (AE) reporting for clinical Quality Program Leads (QPL). However, there were limitations to using solely a machine learning model.

**Objective:** Our primary objective was to propose a robust method to compute the probability of AE underreporting that could complement our machine learning model. Our model was developed to enhance patients safety while reducing the need for on-site and manual QA activities in clinical trials.

**Methods:** We used a Bayesian hierarchical model to estimate the site reporting rates and assess the risk of underreporting. We designed the model with Project Data Sphere clinical trial data that is public and anonymized.

**Results:** We built a model that infers the site reporting behavior from patient-level observations and compares them across a study to enable a robust detection of outliers between clinical sites.

**Conclusion:** The new model will be integrated into the current dashboard designed for clinical Quality Program Leads. This approach reduces the need for on-site audits, shifting focus from source data verification (SDV) to pre-identified, higher risk areas. It will enhance further quality assurance activities for safety reporting from clinical trials and generate quality evidence during pre-approval inspections.

The preprint version of this work is available on MedRxiv: https://doi.org/10.1101/2020.12.18.20245068

**Key points:**

- Safety underreporting is a recurrent issue in clinical trials that can impact patient safety and data integrity
- We used a Bayesian hierarchical model to estimate the site reporting rates and assess the risk of underreporting.
- This model complements our previously published machine learning approach and is used by clinical quality professionals to better detect safety underreporting.

## 1. Introduction

Adverse event (AE) underreporting has been a recurrent issue raised during Health Authorities (HA) Good Clinical Practices (GCP) inspections and audits [1]. Moreover, safety underreporting poses a risk to patient safety and data integrity [2]. The previous clinical Quality Assurance (QA) practices used to detect AE underreporting rely heavily on investigator site and study audits. Yet several sponsors and institutions have had repeated findings related to safety reporting, leading to delays in regulatory submissions.

In a previous project [3], we developed a predictive model that enables pharmaceutical sponsors oversight of AE reporting at the program, study, site and patient level. We validated and reproduced our model using a combination of internal data and an external dataset [4].

While the machine learning model has been successfully implemented since May 2019, there was a need to address calibration robustness. The first model relied on point predictions at the visit level and assumed Poisson distributed residuals. The decision to flag underreporting sites depended on the way these residuals were aggregated and we have been observing instabilities in longer running studies. This motivated us to tackle the problem by the other end and find a model for the distribution of adverse events reported by sites. The biggest value of point estimates from our initial machine learning model is for our stakeholders to direct their investigations, for instance which patient to target during an investigator site audit. On the other hand, an automated underreporting alert relies on well-calibrated probabilities for risk estimate. This was the main motivation for this project. These two solutions will be offered in parallel to quality professionals at our organization, the probabilistic one to quantify the risk of underreporting, and the machine learning based model to provide a basis for in-depth investigations and audits.

The project has been conducted by a team of data scientists, in collaboration with clinical and QA subject matter experts (SMEs). This project was part of a broader initiative of building data-driven solutions for clinical QA to complement and augment traditional QA approaches and to improve the quality and oversight of GCP - and Good Pharmacovigilance Practices (GVP) - regulated activities.

## 2. Methods

### 2.1. Outline

The primary objective was to develop a robust methodology to assess the risk of adverse events underreporting from investigator sites. The scope remained focused on adverse events - not adverse drug reactions-that should occur in clinical trials. Good clinical practice requires all AEs, regardless of a causal relationship between the drug and events, to be reported timely to the sponsor [2]. Underreporting of safety events is a frequent and repetitive issue in clinical trials [1,5] with many consequences, e.g. delayed approval of new drugs [6–7] or amplifying shortcomings of safety data collection in randomized controlled trials [8].

The traditional way to detect AE underreporting in clinical trials is to conduct thorough site audits [9], on top of monitoring activities and through manual SDV. For sponsors with thousands of sites to audit, this is not manually scalable, hence the strong need for a data-driven approach.

Unfortunately, we will never know how many AEs should have been reported, it is something we have to infer from the data. In other words, we are dealing with an unsupervised anomaly detection problem where we do not observe the true labels. The typical way to solve this type of problem is to fit a probability distribution to the data and compare individual data points to that distribution. To do so, one can compute the likelihood of data points under the distribution and flag values below a certain threshold. If the distribution is normal, this is equivalent to flagging points beyond a certain number of standard deviations from the mean. In more general cases, the likelihood is less interpretable, and one might prefer to compute a tail area under the distribution, namely the probability to make an observation at least as extreme as a given data point. The definition of “extreme” will depend on the context and can be adapted to a specific problem. In our case, we can compute the probability that a random site from a given study would have a lower reporting rate than the one under consideration.

In a previous work [3], to infer the distribution of adverse events, we exploited the variety of covariates available at the patient and visit levels to estimate a conditional density *p*(*y*_*visit*_ | *x*_*visit*_;θ) via machine learning. For site-level estimates, we aggregated the visit-level distributions to patient-level and then site-level via successive summations. While this method tracks adverse event data generation at various resolutions, the aggregation introduced biases in the form of systematic over- or underestimation for certain sites, in particular in longer-running trials, probably due to the addition of non-independent errors. As a result, the risk assessment of safety underreporting from investigator sites was not well calibrated. This motivated the top-down approach presented here, as we were ultimately interested in the selection of sites for audits.

To further increase the robustness of the risk assessment, we adopted a Bayesian approach to quantify uncertainties through posterior probability distributions. This is a very appealing property in sectors where risk management is essential such as healthcare or finance. In our case, a clear estimate of the probability of underreporting from the different sites enables targeting of the riskiest sites to audit, and, on the positive side, to gain confidence in the completeness of the collected safety data.

The general methodology of Bayesian data analysis is well described in the literature [10]. The main idea is to build a probabilistic model for the observed data, denoted by *X*, that contains unobserved parameters, collectively denoted by θ. This model relies on a subjective assessment of the distribution of the parameters in the form of a prior distribution *p*(θ), which is more or less sharp depending on the degree of certainty of the prior knowledge. The relation between the parameters and the observed data is expressed by the likelihood function *p*(*X* | θ). The goal of Bayesian inference is to refine the prior distribution, once the data is observed via the application of Bayes’ theorem, and obtain the posterior distribution *p*(θ | *X*) = *p*(*X* | θ)*p*(θ)/*p*(*X*)used to make decisions, estimate parameters, or assess risks. If the observed data is compatible with the prior distribution, the posterior distribution will typically have a smaller spread than the prior. If it is less compatible, then the likelihood and the prior will compete and the posterior distribution will represent a compromise.

In our problem, the observed data is numbers of AEs reported by the sites, grouped by patients, and parameters could be unobserved adverse event reporting rates from the individual sites. We emphasize that there can be several competing models for the observed data, and the goal is to find one that is complex enough to capture the structures of interest, namely safety underreporting in our case, but as simple as possible to speed up computations and convey the clearest insights to stakeholders.

### 2.2. Data

We developed this project on our sponsored clinical trials, but this methodology is applicable to any trial. For illustration, we used public data from the project Data Sphere (PDS), “*an independent, not-for-profit initiative of the CEO Roundtable on Cancer’s Life Sciences Consortium (LSC), operates the Project Data Sphere platform, a free digital library-laboratory that provides one place where the research community can broadly share, integrate and analyze historical, patient-level data from academic and industry phase III cancer clinical trials*” [11]. PDS data was fit for purpose to demonstrate the approach presented in this paper and are publicly available (lifting any concerns for data privacy and security). Specifically, we used the control arm of the registered clinical trial NCT 00617669 [12]. Of note, the data had been further curated to remove duplicate adverse events. The dataset included 468 patients from 125 clinical sites.

From the clinical trial data, we extracted for our analysis the count of adverse events reported by patient, grouped by investigator site (see Table 1).

**Table 1.** Sample of input data (the whole set includes 125 sites and 468 patients)

| Clinical Investigator Site | Count of observed AEs / patient | Number of Patients |
| --- | --- | --- |
| site 3001 | (4, 1) | 2 |
| site 3002 | (2, 2, 1, 2, 5, 5, 5, 1) | 8 |
| site 3003 | (7,4) | 2 |
| site 3004 | (3, 27 ,8) | 3 |
| site 3005 | (12, 6) | 2 |
| site 3006 | (2, 4) | 2 |
| site 3007 | (5) | 1 |
| site 3008 | (11, 4, 16, 31, 23, 23) | 6 |
| site 3009 | (6, 6) | 2 |
| site 3010 | (21, 10, 6, 17, 10, 26, 19, 18, 1, 18, 23,...) | 17 |

### 2.3. Model

We had access to patient level observations, but we needed to make decisions at the site level based on comparisons across the whole study, so a hierarchical model was well indicated as it would capture this three levels structure. Concretely, we assumed that adverse event reporting by a given site could be modelled by a Poisson process. The observed numbers of adverse events for each of the *n*_*i*_ patients reported by the *i*-th site would then be realizations of the corresponding Poisson process,

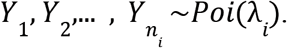

We further assumed that the reporting rates λ _*i*_, *i* = 1, 2, …, *N*_*sites*_ of sites were drawn from a single study-level Gamma distribution Γ(μ_*study*_, σ_*study*_)to model the variability of reporting behaviours among the sites, and we picked vague hyperpriors for the study parameters μ_*study*_~*Exp*(0. 1) and σ_*study*_~*Exp*(0. 1) to account for uncertainty. The parameterization of the Gamma distribution by the mean and standard deviation rather than the more usual shape and rate parameters was intended to make the posterior distribution more interpretable. All these relations are summarized in a graphical representation (Fig. 1). The circles represent random variables, shaded when they correspond to observed data. Arrows indicate conditional dependencies, and plates represent repeated elements, with their labels indicating how many times. The parameters of the hyperprior distributions were chosen so that data simulated by sampling the prior distribution had a similar range as the observed adverse events. We also ran the analysis with wide uniform hyperpriors to check the sensitivity of the inference to the choice of hyperpriors and obtained essentially the same posterior distributions (see the code and the analysis in the Supplementary Material #2).

**Figure 1.**
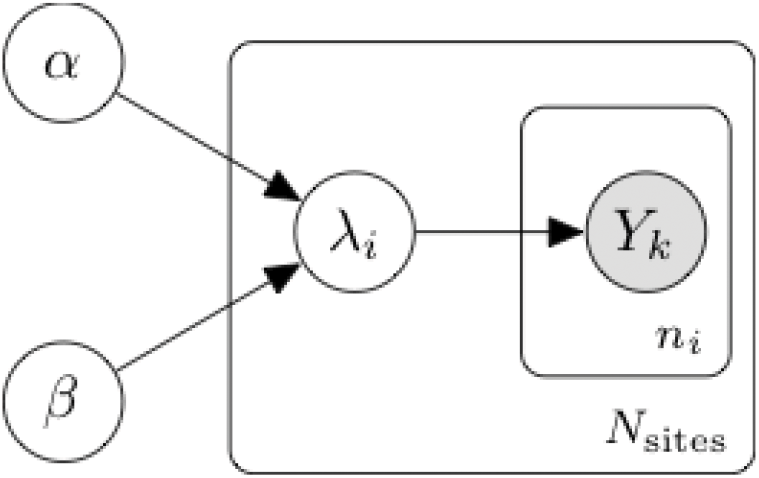
Graphical representation of the adverse event reporting model

In this hierarchical model, the posterior distribution of the site reporting rates λ_*i*_ given the observed numbers of AEs per patient reflects how many AEs per patient we expect to see at the individual sites, and the distribution width indicates the degree of certainty in these estimates. This width typically depends on the number of observations and their spread. Even for sites with less patients, the mechanism of information borrowing enabled by the hierarchical structure leads to more robust estimates of the reporting rates.

The joint posterior distribution of the study level parameters μ_*study*_ and σ_*study*_ characterizes safety reporting patterns of a study and depends on the nature of the disease (e.g. cancers vs. cardiovascular diseases, etc.), the drug mechanism of action, the drug mode of administration, the design and execution of the clinical trial, and so on. The posterior expectation value of μ_*study*_ immediately gives the posterior expectation value of the reporting rate of a site taken at random from that study, and in turn the expected number of adverse events reported by a patient taken at random from that site. The posterior distribution of σ_*study*_ characterizes the variability among the sites of that study. If this analysis is repeated on different studies, the posterior distributions of the parameters μ_*study*_ and σ_*study*_ allow us to compare the reporting patterns of the different studies.

### 2.4 Inference and underreporting detection

Efficient sampling of the posterior distribution of hierarchical models requires specialized methods [13] such as the Hamiltonian Monte Carlo algorithm [14], which is readily implemented in modern probabilistic programming libraries. We used the PyMC3 library [15], and our code is available as a Jupyter notebook [16].

Algorithms of the Markov Chain Monte Carlo family return a sequence of samples of the posterior distribution, in our case a collection of 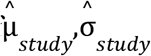 and 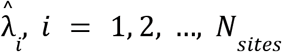. These samples are typically used to compute expectation values with respect to the posterior. We started with the means 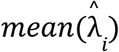 and standard deviations 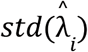 of the site reporting rate samples 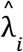 to summarize their distributions, but we were ultimately interested in measuring the risk of underreporting. One natural way to do it is to compute the expected left tail area of the inferred site rates under the posterior (study-level) distribution of reporting rates. This corresponds to the probability that a yet unseen reporting rate drawn randomly from the posterior distribution falls below the inferred site reporting rates. To estimate this posterior probability, for each pair of 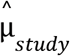 and 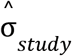 returned in the sample of the Markov chain, we sampled a reference rate 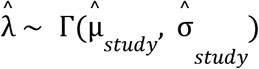 and for each site computed the proportion of samples of the Markov chain such that 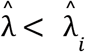 to estimate the rate tail area, 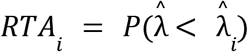.

The output is available in the code repository [16] and a sample of the sites with the top and bottom tail areas is presented in table 2. The inferred values of site 3046 illustrate interesting features of this model. Despite having a single observation of zero reported AEs, the inferred rate is still quite high, driven by information borrowed from the other sites, but with a certain uncertainty, characterized by a higher standard deviation than other sites with low numbers of reported AEs.

**Table 2.** This table displays a sample of the model output with the lowest rate tails areas, together with summary statistics of the inferred AE reporting rates (out of 468 patients in 125 clinical investigator sites). The lowest rate tail areas indicate sites with suspiciously low numbers of reported AEs, and QA activities should be focused on them.

| Clinical Investigator Site | Mean AE rate | Standard Deviation AE rate | Rate tail area | Count of observed AE / patients |
| --- | --- | --- | --- | --- |
| site 3030 | 0.473701 | 0.216417 | 0.00425 | (0, 0, 0, 1, 0, 0, 0, 0, 0, 2) |
| site 3036 | 0.922081 | 0.471779 | 0.01250 | (0, 1, 0, 1) |
| site 3037 | 1.330058 | 0.511819 | 0.02120 | (1, 0, 1, 3, 0) |
| site 3046 | 1.599186 | 1.191997 | 0.03470 | (0) |
| site 3035 | 2.262649 | 1.036725 | 0.05175 | (0, 3) |
| site 3032 | 2.265597 | 1.030830 | 0.05235 | (3, 0) |
| site 3018 | 2.627988 | 0.806107 | 0.06575 | (6, 1, 2, 0) |
| site 3039 | 2.727528 | 1.125527 | 0.06990 | (3, 1) |
| site 3038 | 2.865798 | 0.821121 | 0.07370 | (4, 1, 3, 2) |
| site 3002 | 3.047176 | 0.606155 | 0.08005 | (2, 2, 1, 2, 5, 5, 5, 1) |
| site 3001 | 3.212805 | 1.227723 | 0.08990 | (4, 1) |
| site 3112 | 3.212392 | 1.234220 | 0.09110 | (5, 0) |
| site 3028 | 3.387632 | 1.756067 | 0.09915 | (2) |
| site 3006 | 3.675763 | 1.323994 | 0.10695 | (2, 4) |
| site 3105 | 4.267382 | 1.927275 | 0.13710 | (3) |

When it comes to deciding which sites to flag for under-reporting, a threshold has to be set by quality leads, as the values of the risk metrics cover a wide spectrum displayed in Figure 2.

**Fig. 2.**
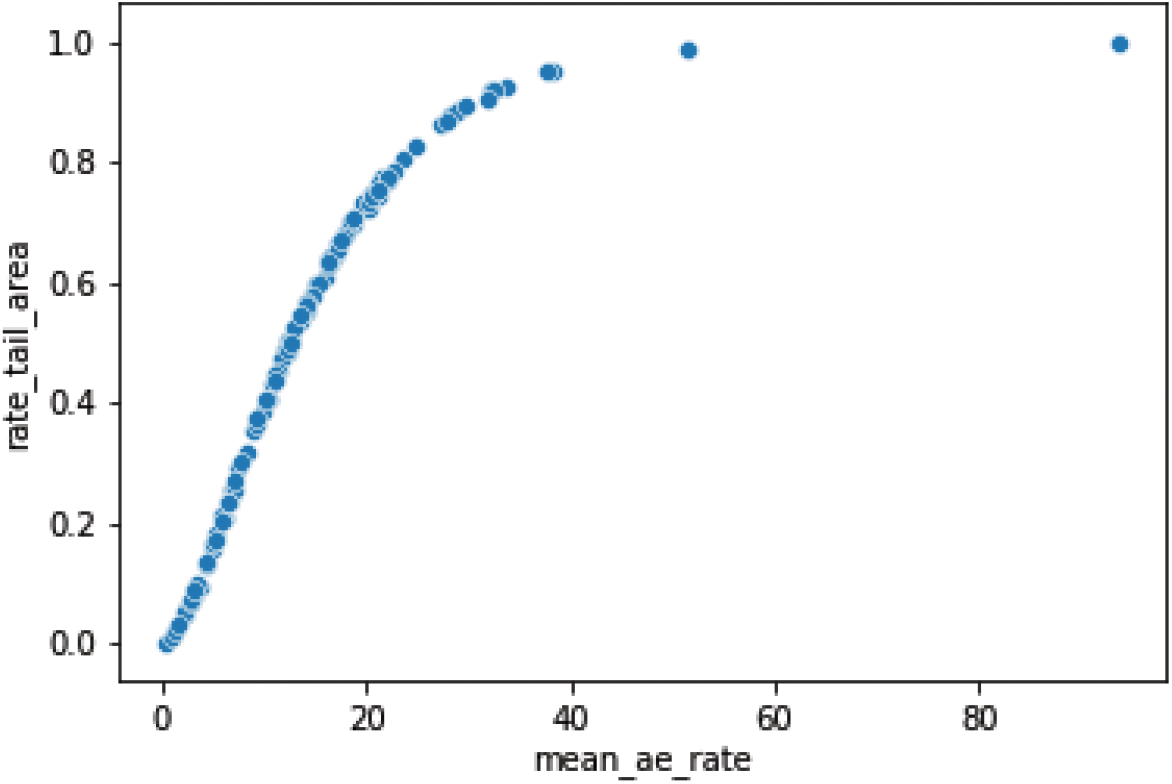
The rate tail area risk metric as a function of the posterior mean site rate.

The relationship between the posterior mean site rates 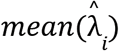 and the corresponding posterior rate tail areas *RTA*_*i*_ follows the cumulative distribution function of the posterior predictive distribution of the reporting rates (Fig. 2). There is no definitive rule to determine how low rate tail areas indicate underreporting. Low values might be due to the inherent variability of safety reporting. Nevertheless, auditing efforts should focus on the lowest values, for instance according to different alert levels at prespecified thresholds, e.g. 0.05 and 0.15, or up to a gap in the distribution of tail areas for more homogeneity in the QA process.

## 3. Discussion

The method presented here is applicable to completed studies to assess which sites might pose a risk of underreporting. In particular, it can demonstrate a degree of certainty in the completeness of collected safety data. In ongoing studies, patients do not enroll all at the same time which introduces more variability in the numbers of reported AEs. The longer a patient has been enrolled, the more AEs have been reported. We can still apply the same method, provided we select as observations the accrued number of adverse events for each patient up to the *n*-th visit and exclude patients who have not reached that milestone. This analysis can be repeated for different values of *n*. In particular, when a study is close to a data-base lock (e.g. before performing an interim analysis), the model could be used to guide quality leads and/or clinical operations staff to detect underreporting sites and trigger queries and AE reconciliation. Hence, this gives reassurance to health authorities inspectors that AE underreporting had been detected, corrective actions had been implemented and integrity of the data had not been compromised [2].

An added benefit of the proposed Bayesian approach and the selected risk metrics is that the outputs are calibrated probabilities. The results of this underreporting risk assessment conducted on different studies and at different milestones are immediately comparable. A sponsor overseeing several studies can thus keep an overview of all of them, and monitor the evolution of the risk of underreporting over time.

Yet this simple Bayesian model ignores the granularity of the available data that goes down to the visit level, the associated time series structure, and a whole collection of covariates that can predict the occurrence of AEs. As mentioned in the methods section, we used this information in our previous work to estimate the number of AEs reported at a single visit, *p*(*y* _*visit*_ | *x*_*visit*_;θ), with machine learning algorithms, but the estimated risks were not well calibrated. Now that we have established that Bayesian methods can address this issue, we plan to explore a middle ground between classical machine learning and probabilistic modeling, namely in the space of Bayesian neural networks, where we can find models that use all covariates but still output calibrated risks of underreporting. This approach will obviously require access to a certain amount of clinical data, which is possible only for a few selected entities such as big clinical trial sponsors, so we still think there is value in the simple approach presented here when it comes to assess individual trials.

In parallel to developing a new model for detection of AE underreporting, we have been piloting a machine learning model with Quality Assurance (QA) staff since May 2019. The outputs of the Bayesian approach will be integrated in the current QA dashboard, together with the outputs of the ML model [3] that are already available to Quality Program Leads. For example, low values of the rate tail areas would indicate sites with suspiciously low numbers of reported AEs, and QA activities should be primarily directed on them. The advantage of combining both approaches is to have AE patient level predictions (from the ML model) and detection of AE underreporting at the site level (using the Bayesian approach). This will enhance further the quality assurance activities for safety reporting from clinical trials.

This model was developed during the Covid-19 pandemic where on-site audits could not be performed [17]. Having a data product enabling remote monitoring of safety reporting from investigator sites was essential to ensure business continuity for clinical quality assurance activities [18]. Our approach has the potential to reduce the need for on-site audits and thereby shift the focus away from source data validation and verification towards pre-identified, higher risk areas. It can contribute to a major shift for QA, where advanced analytics can detect and mitigate issues faster, and ultimately accelerate approval and patient access of innovative drugs.

Jianing Di *et al*. [19] also proposed the use of Bayesian methods for adverse events monitoring with a more clinical purpose. Their approach focused on the continuous monitoring of safety events to address the lack of knowledge of the full safety profile of drugs under clinical investigation. Their model could be applied for signal detection in early-phase trials and could also give further evidence to independent data monitoring committees for late stage studies. We developed a different model as our focus was on sites rather than patients. This illustrates one of the strengths of Bayesian data analysis, where different models of the same data can be optimized to answer different questions about the underlying process.

Clinical trials generate large amounts of data traditionally analysed with frequentist methods, including statistical tests and population parameter estimations, aimed at clinical questions related to efficacy and safety. There has been a push in recent years for Bayesian adaptive designs that have the potential to accelerate and optimize clinical trial execution. Examples include Bayesian sequential design, adaptive randomization, and information borrowing from past trials. For example, in a study redesigning a phase III clinical trial, a Bayesian sequential design could shorten the trial duration by 15 to 40 weeks and recruited 231 to 336 fewer patients [20]. Our approach for the detection of safety underreporting demonstrates the potential of Bayesian data analysis to address secondary questions arising from clinical trials such as quality assurance or trial monitoring. In clinical QA, where the majority of business problems are anomaly detection or risk assessment, there is a good rationale for exploring further applications of Bayesian approaches, for example for identification of laboratory data anomalies in clinical trials or in identifying issues with the number of unreported/reported protocol deviations by clinical study sites.

While the presented method (used together with our machine learning approach [3]) provides a robust strategy to identify AE underreporting, we acknowledge that in rare situations issues could remain hidden. As the majority of activities for clinical trial safety quality oversight have transitioned to be analytics-driven, ad-hoc and on-site quality activities (e.g. clinical investigator site audits) should remain a back-up option for clinical quality assurance organizations.

## 4. Conclusion

In this paper, we presented our approach to quantify the risk of AE underreporting from clinical trial investigator sites. We addressed a shortcoming of the model developed in our previous work that was good at predicting the evolution of safety reporting in clinical studies but failed to properly quantify the probabilities of quality issues.

The new model will be integrated into the current dashboard designed for quality leads. This is part of a broader effort at our Research and Development Quality organization. Similar approaches using statistical modeling and applied to other key risk areas (e.g. informed consent, data integrity) are being developed in order to provide a full set of advanced analytics solutions for clinical quality [3, 4, 17, 21]. We will also continue to explore the application of Bayesian methods to other datasets generated during the conduct of clinical study for QA purposes (e.g. protocol deviations).

However, in order to implement routine, remote, and analytics-driven QA operations, sponsors and agencies will have to continue to collaborate and address challenges such as fit-for-purpose IT infrastructures, automation, cross-company QA data sharing, QA staff data literacy and model validation [17, 21, 22]. While the Covid-19 pandemic accelerated the adoption of new ways of working and pushed innovation further, it also brought new rationales for a change in the QA paradigm, i.e. where advanced analytics can help conducting QA activities remotely, detecting and mitigating issues faster, and ultimately accelerating approval and patient access of innovative drugs.

## Data Availability

The data can be accessed on the Project Data Sphere website.

https://github.com/ybarmaz/bayesian-ae-reporting/blob/main/data.csv

https://data.projectdatasphere.org/projectdatasphere/html/content/104

## Notes

### Author’s contributions

YB developed the model, wrote the code and produced the figures. YB and TM wrote the manuscript. TM quality checked the manuscript. All authors read and approved the final version of the manuscript.

## Acknowledgements

Content review was provided by Kelly Kwon who was employed by Roche/Genentech at the time this research was completed. Project Data Sphere data was curated by Donato Rolo and Björn Koneswarakantha (who were employed by Roche/Genentech at the time this research was completed).

## Compliance with Ethical Standards

### Funding

Funding for development and testing of the safety reporting model was supplied by Roche.

### Ethics Statement

All human subject data used in this analysis were used in a fully de-identified format (see also the link to Project Data Sphere below)

### Data and Code availability

The data can be accessed on Project Data Sphere and through the Supplementary Material #1 https://data.projectdatasphere.org/projectdatasphere/html/content/104

The code and the curated data are available on https://github.com/ybarmaz/bayesian-ae-reporting/

