## Supplementary Material #2 for "Bayesian modeling for the detection of adverse events underreporting in clinical trials"

### Supplementary Materials - Companion notebook

### companion_notebook

May 11, 2021

#### 1 Bayesian modeling for the detection of adverse events underreporting in clinical trials

This is the companion notebook of a paper by Y. Barmaz and T. Ménard on a statistical method for the detection of adverse event underreporting from clinical trial investigator sites.

[1]:
```
import pandas as pd
import numpy as np
import random
import pymc3 as pm
import arviz as az

import seaborn as sns
import matplotlib.pyplot as plt

RANDOM_SEED = 123
```

[2]:
```
data = pd.read_csv('ae_data.csv')
data.head(5)
```

[2]:

| | study | site_number | patnum | ae_count_cumulative |
|---|---|---|---|---|
| 0 | NCT00617669 | 3001 | 16 | 4 |
| 1 | NCT00617669 | 3001 | 456 | 1 |
| 2 | NCT00617669 | 3002 | 171 | 2 |
| 3 | NCT00617669 | 3002 | 248 | 2 |
| 4 | NCT00617669 | 3002 | 304 | 1 |

[3]:
```
# Helper functions and data to visualize AE counts frequencies and their␣
↪predicted values

df_obs = (data
          .groupby('ae_count_cumulative')[['patnum']]
          .count()
          .reset_index()
          .rename({'patnum': 'observed', 'ae_count_cumulative': 'ae_count'},␣
↪axis=1)
          .assign(norm_observed=lambda x: x.observed/x.observed.sum())
```

```
        )

df_obs = (pd.DataFrame({'ae_count': range(50)})
          .merge(df_obs.reset_index(drop=True), how='left')
          .fillna(0)
          .assign(observed=lambda x: x.observed.astype(int))
        )

def format_barplot(ax):
    ax.xaxis.set_major_locator(plt.MultipleLocator(10))
    ax.xaxis.set_minor_locator(plt.MultipleLocator(10))
    ax.xaxis.set_major_formatter(plt.FuncFormatter(lambda x, tick: int(x)))
    return ax

def plot_observed_ae_frequencies():
    ax = format_barplot(sns.barplot(x='ae_count', y='norm_observed',␣
 ↪data=df_obs))
    ax.set_title('Observed AE count frequencies')
    plt.show()

def plot_predicted_ae_frequencies(predicted_checks, prefix=''):
    ae_values, prior_counts =np.unique(predicted_checks['observations'].
 ↪flatten().astype(int), return_counts=True)
    df_prior_obs = df_obs.merge(pd.DataFrame({'ae_count': ae_values,
                                              'prior': prior_counts,
                                              'norm_prior': prior_counts/
 ↪prior_counts.sum()}),
                                how='left')

    ax = format_barplot(sns.barplot(x='ae_count', y='norm_prior',␣
 ↪data=df_prior_obs))
    ax.set_title(prefix + ' predicted AE count frequencies')
    plt.show()
```

[4]:
```
sites = data['site_number'].values
observed_ae = data['ae_count_cumulative'].values

unique_sites, sites_idx = np.unique(sites, return_inverse=True)
```

#### 1.1 Model with exponential priors

The (observed) count of adverse events reported by the $n_i$ patients of site $i$ are modeled with a Poisson distribution, $Y_i \sim \text{Poi}(\lambda_i)$. The $N_{\text{sites}}$ Poisson rates $\lambda_i$ are in turn modeled as realizations of a random variable unique to the whole study with Gamma distribution $\Gamma(\mu, \sigma)$. The parameters $\mu$ and $\sigma$ are unknown, so we assume a vague prior for both of them, $\mu \sim \text{Exp}(0.1)$ and $\sigma \sim \text{Exp}(0.1)$.

The full joint distribution

$$P(\mu, \sigma, \lambda_i, Y_{i,j}) = P(\mu)P(\sigma) \prod_{i=1}^{N_{\text{sites}}} P(\lambda_i|\mu, \sigma) \prod_{j=1}^{n_i} P(Y_{i,j}|\lambda_i)$$

is coded in the following PyMC3 model:

[41]:
```
with pm.Model() as model:
    mu = pm.Exponential("mu", lam=0.1)
    sigma = pm.Exponential("sigma", lam=0.1)

    rates = pm.Gamma("rates", mu=mu, sigma=sigma, shape=unique_sites.shape)
    observations = pm.Poisson("observations", mu=rates[sites_idx],␣
↪observed=observed_ae)

    # sample a reference rate from the study-level distribution and use it to␣
↪estimate
    # the tail area of the inferred site rates:
    reference_rate = pm.Gamma("reference_rate", mu=mu, sigma=sigma)
    underreporting = pm.Deterministic("underreporting", reference_rate < rates)

    prior_checks = pm.sample_prior_predictive(samples=50,␣
↪random_seed=RANDOM_SEED)
```

```
/Users/barmazy/opt/anaconda3/lib/python3.7/site-
packages/pymc3/distributions/transforms.py:221: RuntimeWarning: divide by zero
encountered in log
  return np.log(x)
```

##### 1.1.1 Prior predictive check

The validity of the hyperprior parameters can be verified with a sample of AE counts from the prior distribution:

[42]:
```
plot_observed_ae_frequencies()
```

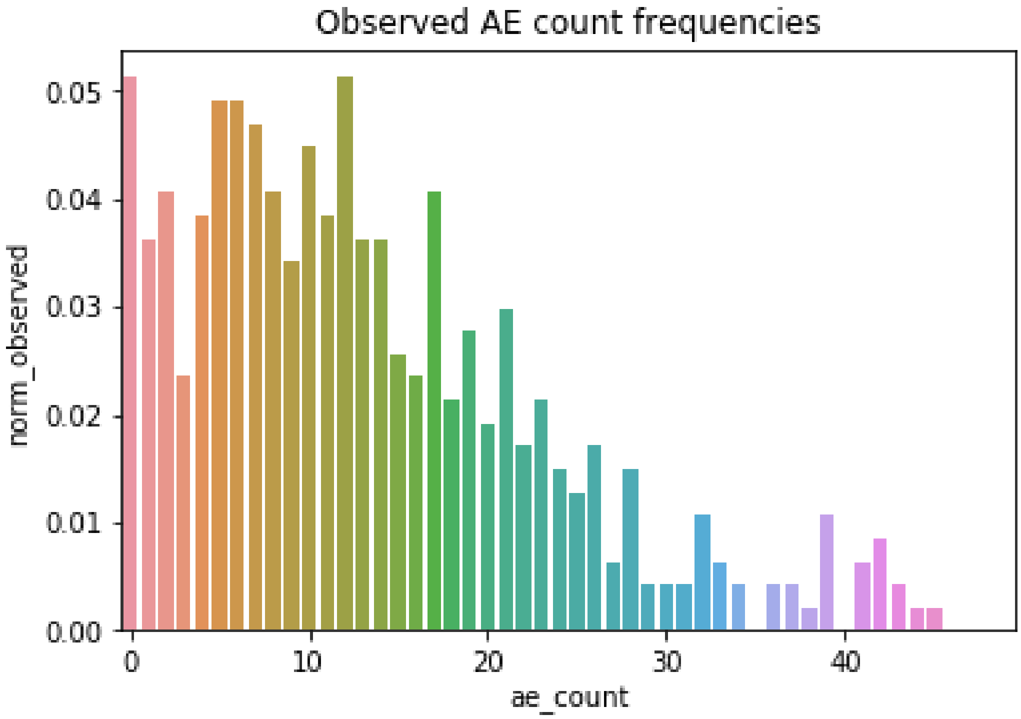


[43]:
```
plot_predicted_ae_frequencies(prior_checks, "Prior")
```

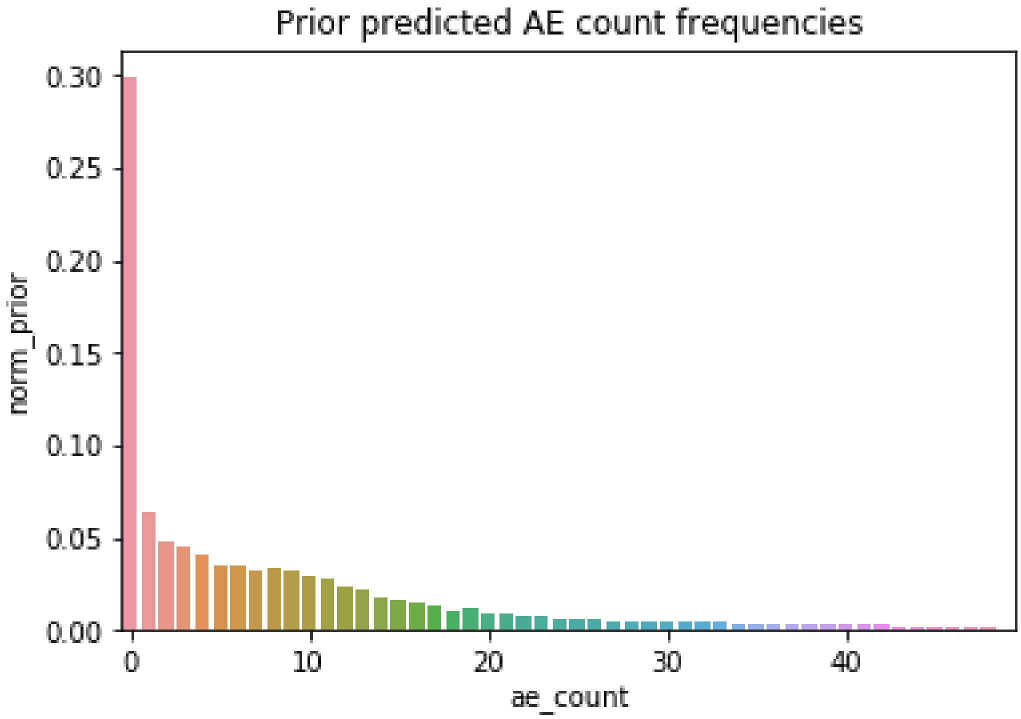

##### 1.1.2 MCMC inference

[44]:
```
with model:
    idata = pm.sample(5000, return_inferencedata=True, random_seed=RANDOM_SEED)
```

```
Auto-assigning NUTS sampler…
Initializing NUTS using jitter+adapt_diag…
Multiprocess sampling (4 chains in 4 jobs)
NUTS: [reference_rate, rates, sigma, mu]
```

```
<IPython.core.display.HTML object>
```

```
Sampling 4 chains for 1_000 tune and 5_000 draw iterations (4_000 + 20_000 draws
total) took 34 seconds.
```

##### 1.1.3 Convergence assessment

[45]:
```
pm.plot_trace(idata, var_names=["mu", "sigma", "rates"], coords={"rates_dim_0":␣
 ↪[0, 1, 2, 3]})
plt.show()
```

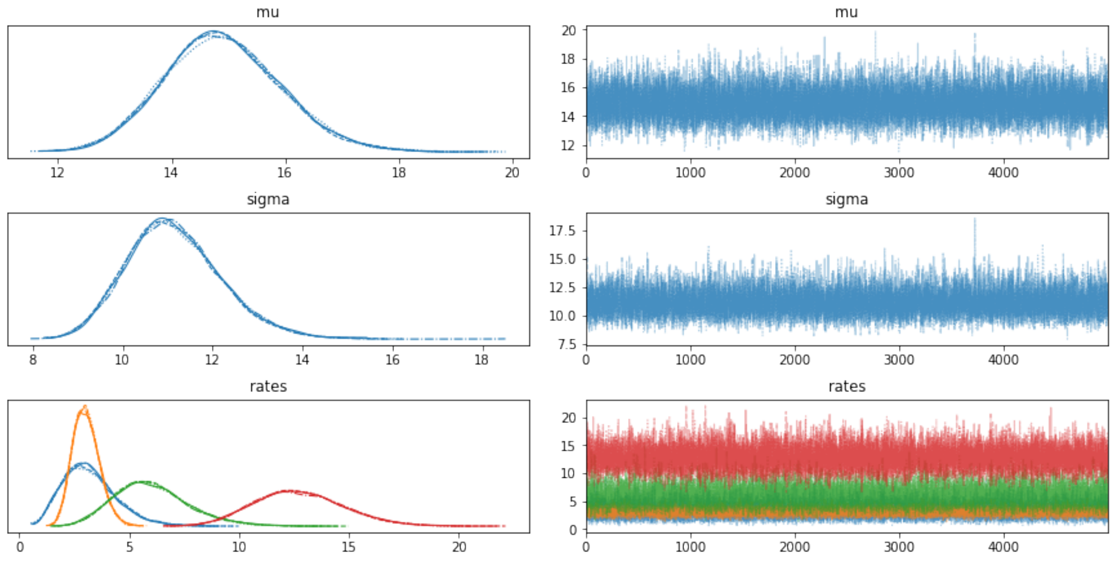


[46]:
```
az.summary(idata, var_names=["mu", "sigma", "rates"], round_to=2).head(n=10)
```

[46]:

| | mean | sd | hdi_3% | hdi_97% | mcse_mean | mcse_sd | ess_bulk | \ |
|---|---|---|---|---|---|---|---|---|
| mu | 14.89 | 1.02 | 12.97 | 16.78 | 0.01 | 0.01 | 19264.06 | |
| sigma | 11.14 | 1.05 | 9.25 | 13.15 | 0.01 | 0.01 | 17569.88 | |
| rates[0] | 3.21 | 1.23 | 1.06 | 5.43 | 0.01 | 0.01 | 32322.59 | |

```
rates[1]   3.05  0.61    1.94     4.19       0.00      0.00  29834.96
rates[2]   6.03  1.69    2.92     9.14       0.01      0.01  27043.80
rates[3]  12.73  2.03    9.04    16.56       0.01      0.01  32704.83
rates[4]   9.35  2.08    5.59    13.32       0.01      0.01  33637.81
rates[5]   3.68  1.32    1.42     6.14       0.01      0.01  31005.83
rates[6]   6.06  2.34    2.17    10.46       0.01      0.01  33462.62
rates[7]  17.93  1.70   14.81    21.17       0.01      0.01  39265.76

          ess_tail  r_hat
mu        16036.76    1.0
sigma     15025.40    1.0
rates[0]  13430.62    1.0
rates[1]  13872.00    1.0
rates[2]  12263.17    1.0
rates[3]  14668.23    1.0
rates[4]  13031.25    1.0
rates[5]  13793.89    1.0
rates[6]  13071.82    1.0
rates[7]  14502.75    1.0
```

##### 1.1.4 Posterior predictive check

[47]:
```
with model:
    ppc = pm.sample_posterior_predictive(idata, var_names=['rates',␣
↪'observations', 'reference_rate', 'underreporting'], random_seed=RANDOM_SEED)

plot_predicted_ae_frequencies(ppc, "Posterior")
```

```
<IPython.core.display.HTML object>
```

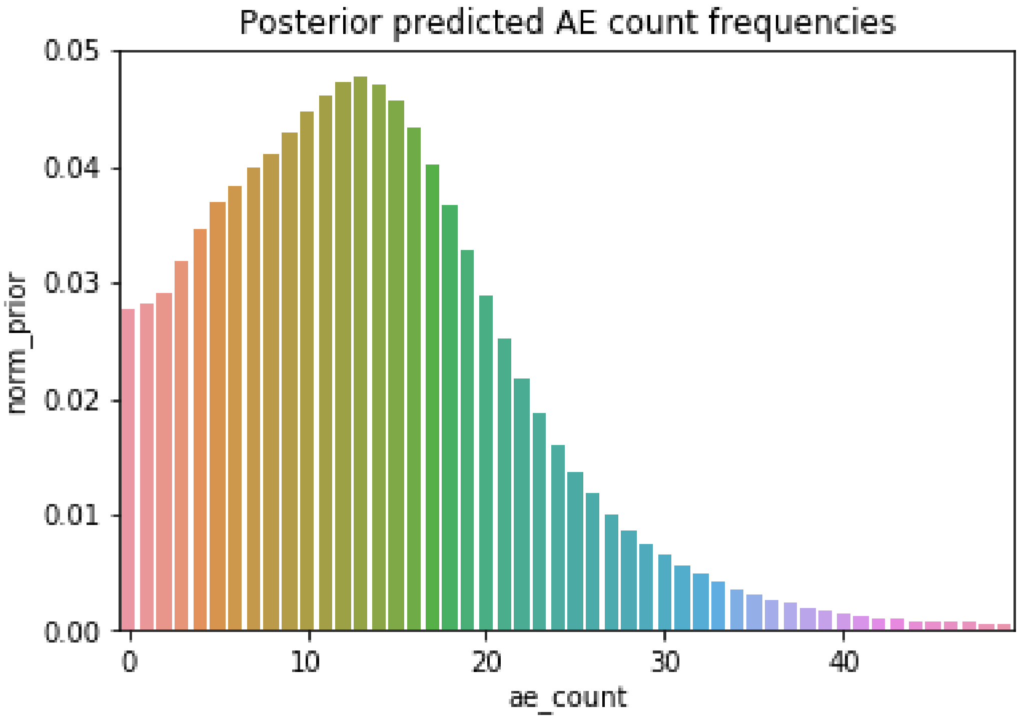


[48]:
```
def summarize_results(idata, unique_sites=unique_sites):
    summary_df = pd.DataFrame({'site': unique_sites,
                               'mean_ae_rate': idata.posterior['rates'].data.
 ↪mean(axis=(0, 1)),
                               'std_ae_rate': idata.posterior['rates'].data.
 ↪std(axis=(0, 1)),
                               'rate_tail_area': idata.
 ↪posterior['underreporting'].data.mean(axis=(0,1))
                              }).merge(pd.DataFrame(data.
 ↪groupby('site_number')['ae_count_cumulative'].apply(list)),
                                       left_on='site',
                                       right_index=True,
                                       how='left'
                                      )
    return summary_df
```

#### 1.2 Underreporting evaluation

The following table summarizes the posterior mean and standard deviation of the site reporting rates as well as their rate tail area under the posterior study-level distribution:

[49]:
```
summarize_results(idata).sort_values('rate_tail_area').head(20)
```

[49]:

```
     site  mean_ae_rate  std_ae_rate  rate_tail_area  \
29   3030      0.473701     0.216417         0.00425
35   3036      0.922081     0.471779         0.01250
36   3037      1.330058     0.511819         0.02120
45   3046      1.599186     1.191997         0.03470
34   3035      2.262649     1.036725         0.05175
31   3032      2.265597     1.030830         0.05235
17   3018      2.627988     0.806107         0.06575
38   3039      2.727528     1.125527         0.06990
37   3038      2.865798     0.821121         0.07370
1    3002      3.047176     0.606155         0.08005
0    3001      3.212805     1.227723         0.08990
111  3112      3.212392     1.234220         0.09110
27   3028      3.387632     1.756067         0.09915
5    3006      3.675763     1.323994         0.10695
104  3105      4.267382     1.927275         0.13710
103  3104      4.300125     1.974931         0.13850
24   3025      4.855567     0.879657         0.16250
33   3034      5.026851     0.897601         0.17070
109  3110      5.182283     0.648874         0.17720
18   3019      5.186425     2.147535         0.18105

                          ae_count_cumulative
29             [0, 0, 0, 1, 0, 0, 0, 0, 0, 2]
35                               [0, 1, 0, 1]
36                            [1, 0, 1, 3, 0]
45                                        [0]
34                                     [0, 3]
31                                     [3, 0]
17                               [6, 1, 2, 0]
38                                     [3, 1]
37                               [4, 1, 3, 2]
1                    [2, 2, 1, 2, 5, 5, 5, 1]
0                                      [4, 1]
111                                    [5, 0]
27                                        [2]
5                                      [2, 4]
104                                       [3]
103                                       [3]
24                         [5, 1, 5, 6, 6, 5]
33                         [8, 3, 6, 2, 3, 7]
109  [7, 0, 2, 10, 8, 7, 10, 5, 2, 4, 2, 4]
18                                        [4]
```

[50]:

```
summarize_results(idata).sort_values('rate_tail_area').tail(20)
```

[50]:
```
      site  mean_ae_rate  std_ae_rate  rate_tail_area  \
70    3071     22.148269     4.437367         0.77430
11    3012     21.667842     1.878398         0.77810
121   3122     22.069505     3.247750         0.77815
112   3113     23.045128     4.534650         0.79050
54    3055     23.462254     3.340926         0.80375
61    3062     24.925184     2.436561         0.83375
60    3061     27.269246     3.592407         0.86350
96    3097     27.737923     3.633415         0.86845
46    3047     28.317340     1.659186         0.88060
67    3068     28.656487     3.690326         0.88150
10    3011     29.058174     2.652968         0.88785
92    3093     29.628805     3.740776         0.89190
122   3123     31.904476     5.324882         0.90480
108   3109     32.397402     3.886327         0.91640
116   3117     32.445790     3.944280         0.91860
53    3054     33.781410     5.499409         0.92435
81    3082     37.605554     4.221412         0.95125
55    3056     38.220139     5.831812         0.95130
105   3106     51.380775     3.494152         0.98725
98    3099     93.595338     6.641897         0.99975

                          ae_count_cumulative
70                                       [23]
11                   [24, 31, 26, 20, 15, 15]
121                                  [44, 1]
112                                      [24]
54                                   [25, 23]
61                           [37, 32, 12, 20]
60                                   [42, 14]
96                                   [39, 18]
46    [28, 42, 39, 32, 4, 32, 21, 30, 15, 42]
67                                   [32, 27]
10                           [20, 28, 41, 29]
92                                   [39, 22]
122                                      [34]
108                                  [42, 25]
116                                  [24, 43]
53                                       [36]
81                                   [45, 33]
55                                       [41]
105                          [17, 65, 75, 53]
98                                  [140, 57]
```

[32]:
```
ax=sns.scatterplot(x='mean_ae_rate', y='rate_tail_area',␣
 ↪data=summarize_results(idata))
plt.show()
```

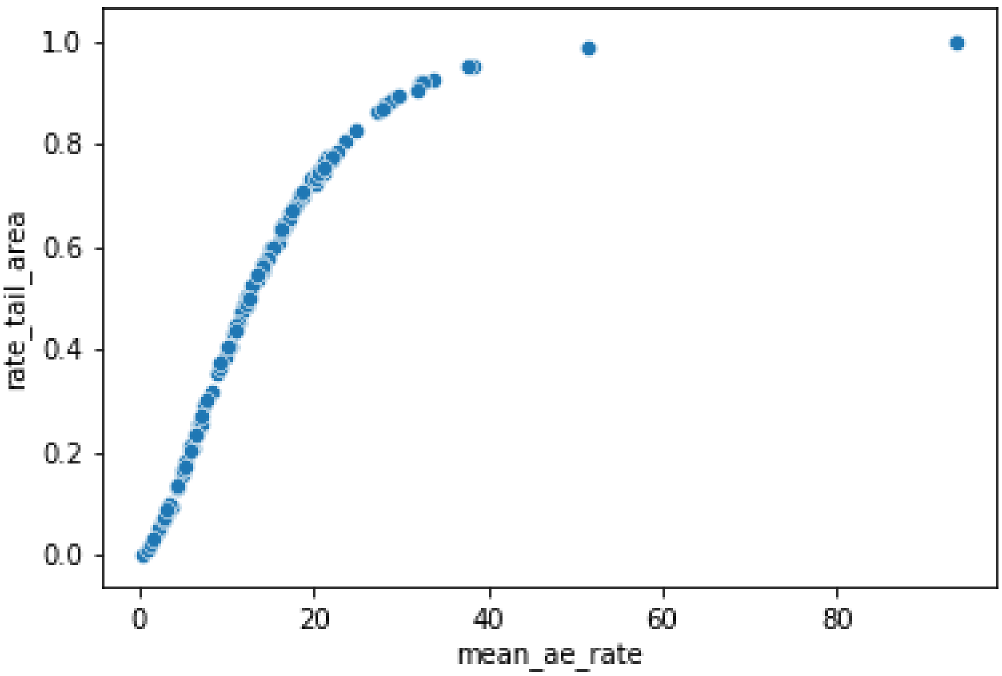


##### 1.3 Model with uniform priors

To check sensitivity to the choice of priors, we can run the same analysis with wide uniform priors $\mu \sim U(0, 50)$ and $\sigma \sim U(0, 50)$:

[14]:
```
with pm.Model() as model_uniform:
    mu = pm.Uniform("mu", lower=0, upper=50)
    sigma = pm.Uniform("sigma", lower=0, upper=50)

    rates = pm.Gamma("rates", mu=mu, sigma=sigma, shape=unique_sites.shape)
    observations = pm.Poisson("observations", mu=rates[sites_idx],␣
 ↪observed=observed_ae)

    reference_rate = pm.Gamma("reference_rate", mu=mu, sigma=sigma)
    underreporting = pm.Deterministic("underreporting", reference_rate < rates)

    prior_checks = pm.sample_prior_predictive(samples=50,␣
 ↪random_seed=RANDOM_SEED)
```

[15]:
```
plot_predicted_ae_frequencies(prior_checks, "Prior")
```

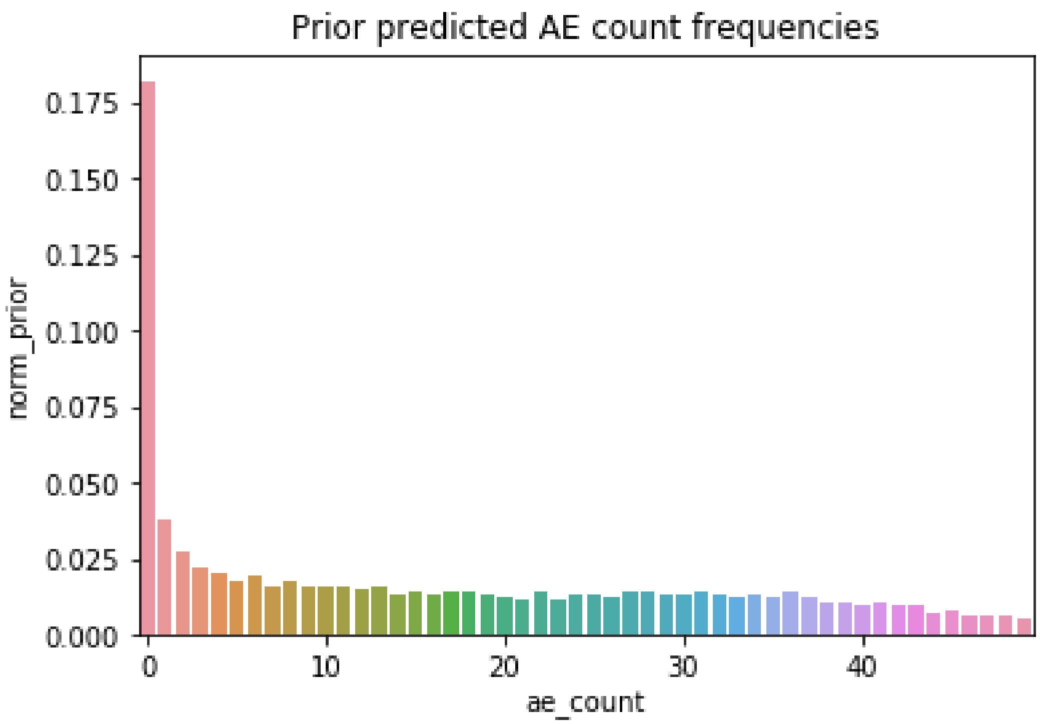


[16]:
```
with model_uniform:
    idata_uniform = pm.sample(1000, return_inferencedata=True,␣
 ↪random_seed=RANDOM_SEED)
```

```
Auto-assigning NUTS sampler…
Initializing NUTS using jitter+adapt_diag…
Multiprocess sampling (4 chains in 4 jobs)
NUTS: [reference_rate, rates, sigma, mu]
```

```
<IPython.core.display.HTML object>
```

```
Sampling 4 chains for 1_000 tune and 1_000 draw iterations (4_000 + 4_000 draws
total) took 20 seconds.
```

[17]:
```
pm.plot_trace(idata_uniform, var_names=["mu", "sigma", "rates"],␣
 ↪coords={"rates_dim_0": [0, 1, 2, 3]})
plt.show()
```

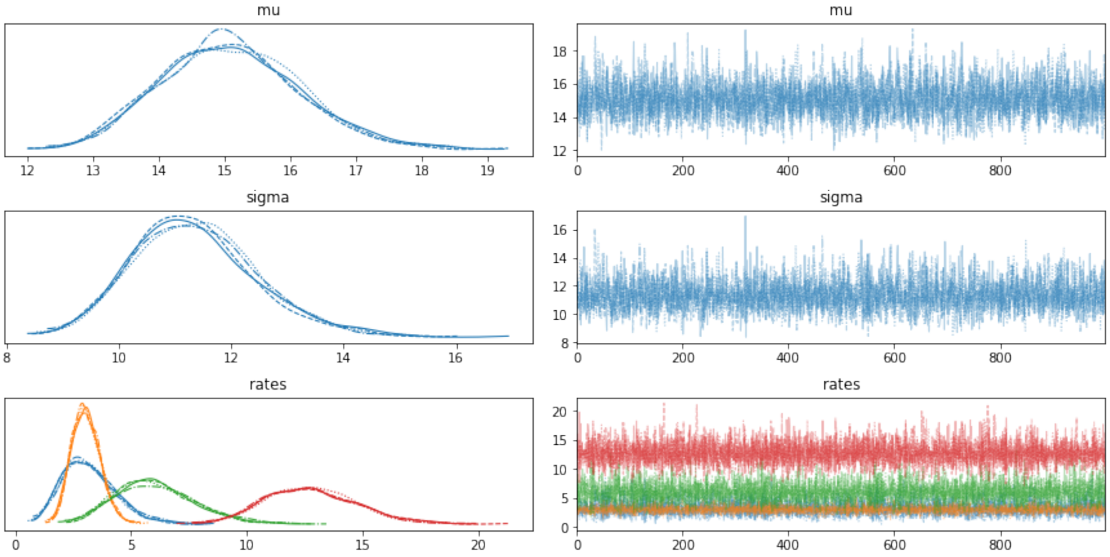


[18]:
```python
az.summary(idata_uniform, var_names=["mu", "sigma", "rates"], round_to=2).
↪head(n=10)
```

[18]:
```
           mean    sd  hdi_3%  hdi_97%  mcse_mean  mcse_sd  ess_bulk  \
mu        15.08  1.06   13.13    17.04       0.02     0.01   3362.49
sigma     11.35  1.10    9.41    13.52       0.02     0.01   2973.48
rates[0]   3.19  1.18    1.18     5.37       0.02     0.01   5489.27
rates[1]   3.05  0.62    1.89     4.17       0.01     0.01   6508.06
rates[2]   6.04  1.71    3.11     9.34       0.02     0.02   6190.23
rates[3]  12.78  1.98    9.10    16.46       0.02     0.02   8038.37
rates[4]   9.30  2.02    5.60    13.08       0.03     0.02   5921.57
rates[5]   3.65  1.33    1.27     6.05       0.02     0.01   5839.29
rates[6]   6.05  2.28    2.18    10.23       0.03     0.02   5664.67
rates[7]  17.99  1.71   14.98    21.38       0.02     0.02   6711.12

          ess_tail  r_hat
mu         2896.64    1.0
sigma      2922.29    1.0
rates[0]   2706.78    1.0
rates[1]   2521.45    1.0
rates[2]   2755.73    1.0
rates[3]   2610.53    1.0
rates[4]   3255.18    1.0
rates[5]   2488.59    1.0
rates[6]   2549.52    1.0
rates[7]   2664.87    1.0
```

[19]:
```
with model_uniform:
    ppc_uniform = pm.sample_posterior_predictive(idata_uniform,
                                                  var_names=['rates',␣
 ↪'observations', 'reference_rate', 'underreporting'],
                                                  random_seed=RANDOM_SEED)
```

<IPython.core.display.HTML object>

[20]:
```
plot_predicted_ae_frequencies(ppc_uniform, "Posterior")
```

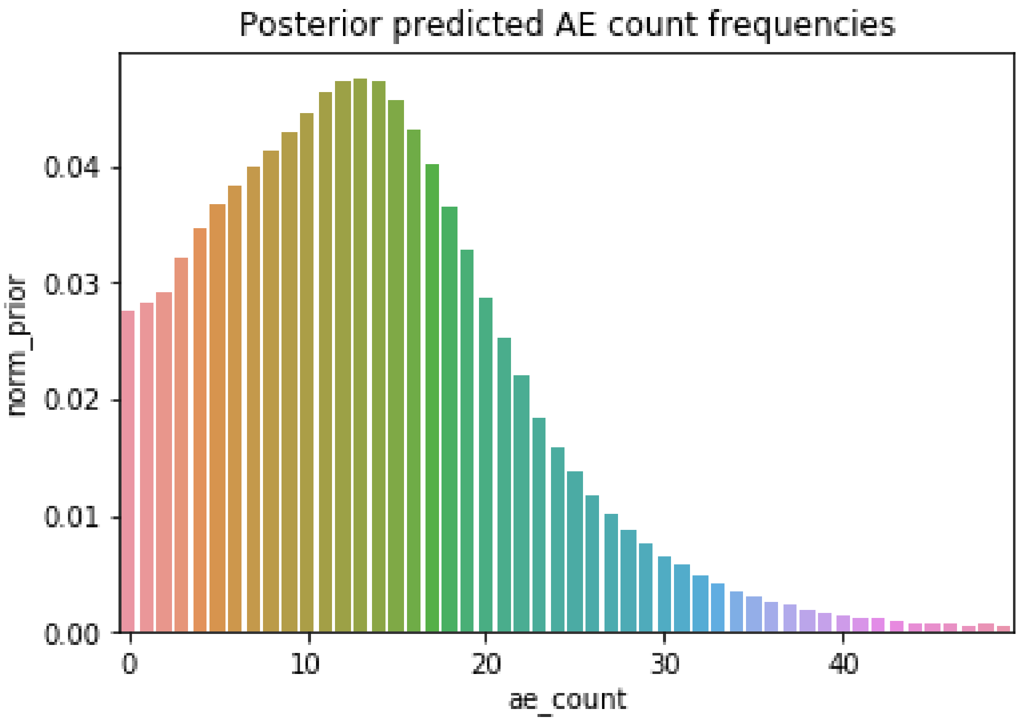


[35]:
```
summarize_results(idata_uniform).sort_values('rate_tail_area').head(20)
```

[35]:

| | site | mean_ae_rate | std_ae_rate | rate_tail_area \ |
|---|---|---|---|---|
| 29 | 3030 | 0.477346 | 0.217253 | 0.00300 |
| 35 | 3036 | 0.920242 | 0.462205 | 0.01200 |
| 36 | 3037 | 1.317767 | 0.502185 | 0.02575 |
| 45 | 3046 | 1.609550 | 1.220536 | 0.03850 |
| 34 | 3035 | 2.274671 | 1.039737 | 0.05650 |
| 31 | 3032 | 2.258359 | 1.027598 | 0.05675 |
| 17 | 3018 | 2.627619 | 0.832237 | 0.06975 |
| 38 | 3039 | 2.734650 | 1.125781 | 0.07625 |
| 37 | 3038 | 2.859058 | 0.828867 | 0.08175 |
| 1 | 3002 | 3.051233 | 0.618543 | 0.08600 |

```
0    3001      3.192507     1.179046         0.08800
111  3112      3.218817     1.197078         0.08950
27   3028      3.396970     1.783684         0.11225
5    3006      3.654133     1.330591         0.11550
103  3104      4.289373     1.976724         0.14375
104  3105      4.286954     1.986052         0.15025
24   3025      4.860359     0.918460         0.16375
33   3034      5.041523     0.913894         0.18125
109  3110      5.176694     0.668167         0.18200
18   3019      5.177377     2.175958         0.18575

                       ae_count_cumulative
29             [0, 0, 0, 1, 0, 0, 0, 0, 0, 2]
35                               [0, 1, 0, 1]
36                            [1, 0, 1, 3, 0]
45                                        [0]
34                                     [0, 3]
31                                     [3, 0]
17                               [6, 1, 2, 0]
38                                     [3, 1]
37                               [4, 1, 3, 2]
1                    [2, 2, 1, 2, 5, 5, 5, 1]
0                                      [4, 1]
111                                    [5, 0]
27                                        [2]
5                                      [2, 4]
103                                       [3]
104                                       [3]
24                         [5, 1, 5, 6, 6, 5]
33                         [8, 3, 6, 2, 3, 7]
109  [7, 0, 2, 10, 8, 7, 10, 5, 2, 4, 2, 4]
18                                        [4]
```

#### 1.4 Smaller trial

To see how this approach performs on smaller trials, we can retain only a fraction of the sites from the full dataset and repeat the analysis.

[39]:
```python
small_sites = unique_sites[:10]

small_data = data[data.site_number.isin(small_sites)]

small_sites = small_data['site_number'].values
small_observed_ae = small_data['ae_count_cumulative'].values
small_unique_sites, small_sites_idx = np.unique(small_sites,␣
 ↪return_inverse=True)
```

[40]:
```
len(small_data)
```

[40]: 45

[23]:
```
with pm.Model() as small_model:
    mu = pm.Exponential("mu", lam=0.1)
    sigma = pm.Exponential("sigma", lam=0.1)

    rates = pm.Gamma("rates", mu=mu, sigma=sigma, shape=small_unique_sites.
 ↪shape)
    observations = pm.Poisson("observations", mu=rates[small_sites_idx],␣
 ↪observed=small_observed_ae)

    reference_rate = pm.Gamma("reference_rate", mu=mu, sigma=sigma)
    underreporting = pm.Deterministic("underreporting", reference_rate < rates)

    prior_checks = pm.sample_prior_predictive(samples=50,␣
 ↪random_seed=RANDOM_SEED)
```

```
/Users/barmazy/opt/anaconda3/lib/python3.7/site-
packages/pymc3/distributions/transforms.py:221: RuntimeWarning: divide by zero
encountered in log
  return np.log(x)
```

[24]:
```
with small_model:
    small_idata = pm.sample(1000, return_inferencedata=True,␣
 ↪random_seed=RANDOM_SEED)
```

```
Auto-assigning NUTS sampler…
Initializing NUTS using jitter+adapt_diag…
Multiprocess sampling (4 chains in 4 jobs)
NUTS: [reference_rate, rates, sigma, mu]
```

```
<IPython.core.display.HTML object>
```

```
Sampling 4 chains for 1_000 tune and 1_000 draw iterations (4_000 + 4_000 draws
total) took 17 seconds.
```

[25]:
```
pm.plot_trace(small_idata, var_names=["mu", "sigma", "rates"],␣
 ↪coords={"rates_dim_0": [0, 1, 2, 3]})
plt.show()
```

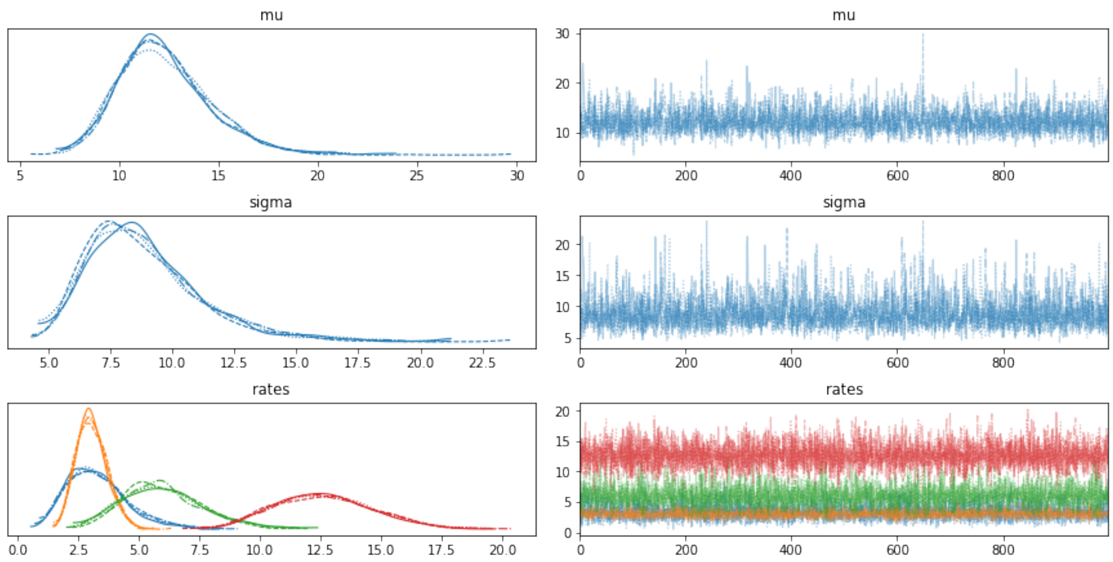


[26]:
```
az.summary(small_idata, var_names=["mu", "sigma", "rates"], round_to=2).
↪head(n=10)
```

[26]:

| | mean | sd | hdi_3% | hdi_97% | mcse_mean | mcse_sd | ess_bulk | \ |
|---|---|---|---|---|---|---|---|---|
| mu | 12.30 | 2.33 | 8.17 | 16.52 | 0.04 | 0.03 | 3498.90 | |
| sigma | 8.90 | 2.47 | 4.98 | 13.35 | 0.05 | 0.03 | 3487.99 | |
| rates[0] | 3.29 | 1.25 | 1.19 | 5.68 | 0.02 | 0.01 | 6270.17 | |
| rates[1] | 3.07 | 0.63 | 1.97 | 4.27 | 0.01 | 0.01 | 6820.20 | |
| rates[2] | 6.00 | 1.68 | 3.04 | 9.26 | 0.02 | 0.02 | 6424.81 | |
| rates[3] | 12.65 | 2.03 | 8.99 | 16.34 | 0.02 | 0.02 | 7567.15 | |
| rates[4] | 9.25 | 2.06 | 5.74 | 13.21 | 0.02 | 0.02 | 7471.45 | |
| rates[5] | 3.74 | 1.38 | 1.39 | 6.29 | 0.02 | 0.01 | 6232.50 | |
| rates[6] | 6.05 | 2.25 | 2.05 | 10.13 | 0.03 | 0.02 | 6729.65 | |
| rates[7] | 17.80 | 1.70 | 14.68 | 20.90 | 0.02 | 0.02 | 6430.40 | |

| | ess_tail | r_hat |
|---|---|---|
| mu | 2862.39 | 1.0 |
| sigma | 2798.98 | 1.0 |
| rates[0] | 2969.51 | 1.0 |
| rates[1] | 2711.71 | 1.0 |
| rates[2] | 2750.70 | 1.0 |
| rates[3] | 3152.18 | 1.0 |
| rates[4] | 3065.36 | 1.0 |
| rates[5] | 3047.52 | 1.0 |
| rates[6] | 2554.36 | 1.0 |
| rates[7] | 2973.06 | 1.0 |

[27]:
```
with small_model:
    small_ppc = pm.sample_posterior_predictive(small_idata,
                                               var_names=['rates',␣
↪'observations', 'reference_rate', 'underreporting'],
                                               random_seed=RANDOM_SEED)

plot_predicted_ae_frequencies(small_ppc, "Posterior")
```

<IPython.core.display.HTML object>

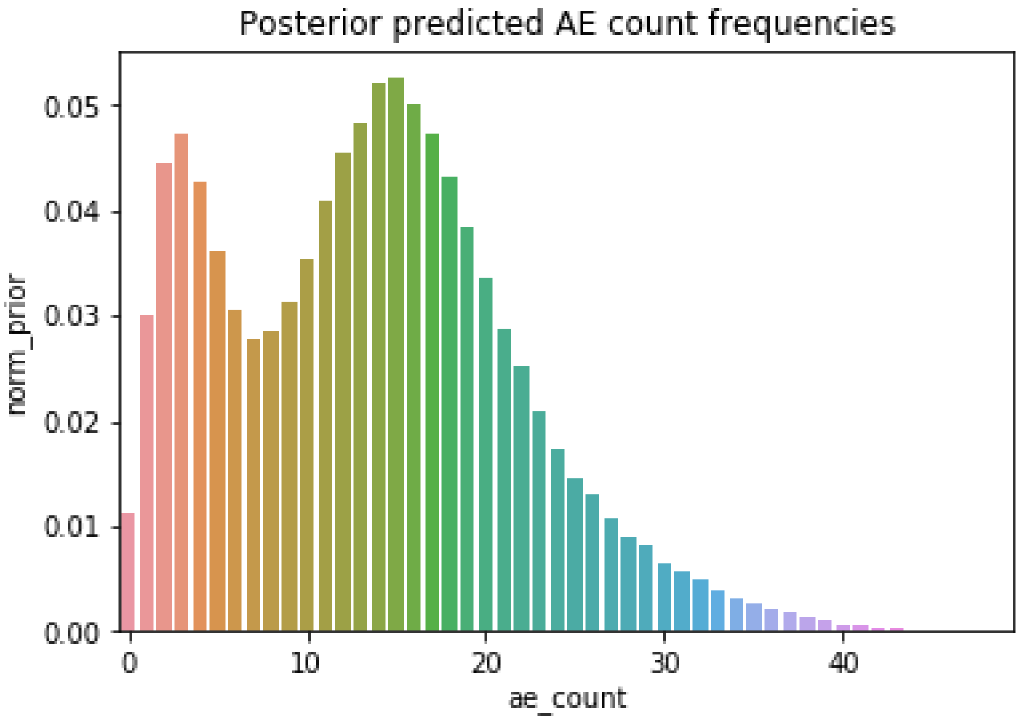


###### 1.4.1 Comparison with the results obtained from the whole dataset:

[28]:
```
df_comp = (summarize_results(small_idata, unique_sites=small_unique_sites)
           .sort_values('rate_tail_area')
           .merge(summarize_results(idata), on='site', how='left',␣
↪suffixes=(None, '_ref'))
           .drop('ae_count_cumulative_ref', axis=1)
          )

df_comp
```

[28]:

| | site | mean_ae_rate | std_ae_rate | rate_tail_area | \ |
|---|---|---|---|---|---|
| 0 | 3002 | 3.073764 | 0.629972 | 0.11000 | |
| 1 | 3001 | 3.287786 | 1.247203 | 0.12650 | |
| 2 | 3006 | 3.738658 | 1.378297 | 0.14475 | |
| 3 | 3003 | 5.999460 | 1.678981 | 0.27125 | |
| 4 | 3007 | 6.047184 | 2.252466 | 0.27250 | |
| 5 | 3009 | 6.474087 | 1.703627 | 0.30425 | |
| 6 | 3005 | 9.253716 | 2.063494 | 0.44550 | |
| 7 | 3015 | 10.602643 | 2.180736 | 0.51475 | |
| 8 | 3004 | 12.645778 | 2.029384 | 0.60450 | |
| 9 | 3010 | 14.674560 | 0.918215 | 0.68500 | |
| 10 | 3014 | 16.318860 | 1.972295 | 0.73400 | |
| 11 | 3008 | 17.796655 | 1.704741 | 0.78100 | |
| 12 | 3013 | 19.326481 | 2.981730 | 0.80525 | |
| 13 | 3012 | 21.549531 | 1.823395 | 0.85350 | |
| 14 | 3011 | 28.706355 | 2.626204 | 0.93450 | |

| | ae_count_cumulative | mean_ae_rate_ref | \ |
|---|---|---|---|
| 0 | [2, 2, 1, 2, 5, 5, 5, 1] | 3.044453 | |
| 1 | [4, 1] | 3.209663 | |
| 2 | [2, 4] | 3.649618 | |
| 3 | [7, 4] | 6.048335 | |
| 4 | [5] | 6.071747 | |
| 5 | [6, 6] | 6.516165 | |
| 6 | [12, 6] | 9.356922 | |
| 7 | [7, 14] | 10.724011 | |
| 8 | [3, 27, 8] | 12.759125 | |
| 9 | [21, 10, 6, 17, 10, 7, 26, 19, 18, 1, 18, 23, … | 14.704852 | |
| 10 | [14, 11, 13, 28] | 16.416935 | |
| 11 | [11, 4, 16, 31, 23, 23] | 17.897772 | |
| 12 | [23, 17] | 19.662940 | |
| 13 | [24, 31, 26, 20, 15, 15] | 21.623752 | |
| 14 | [20, 28, 41, 29] | 29.088904 | |

| | std_ae_rate_ref | rate_tail_area_ref |
|---|---|---|
| 0 | 0.619113 | 0.07650 |
| 1 | 1.247433 | 0.08800 |
| 2 | 1.308859 | 0.09675 |
| 3 | 1.672531 | 0.21575 |
| 4 | 2.289255 | 0.20700 |
| 5 | 1.749248 | 0.23550 |
| 6 | 2.038170 | 0.36875 |
| 7 | 2.225989 | 0.43200 |
| 8 | 1.964344 | 0.50825 |
| 9 | 0.932950 | 0.58450 |
| 10 | 1.944657 | 0.63425 |
| 11 | 1.670321 | 0.68050 |

| | | |
|---|---|---|
| 12 | 2.977025 | 0.72300 |
| 13 | 1.890747 | 0.77750 |
| 14 | 2.728231 | 0.88700 |

[29]:
```
sns.histplot(x=small_idata.posterior['reference_rate'].data.flatten())
plt.title('Posterior distribution of reporting rates from the small dataset')
plt.show()
```

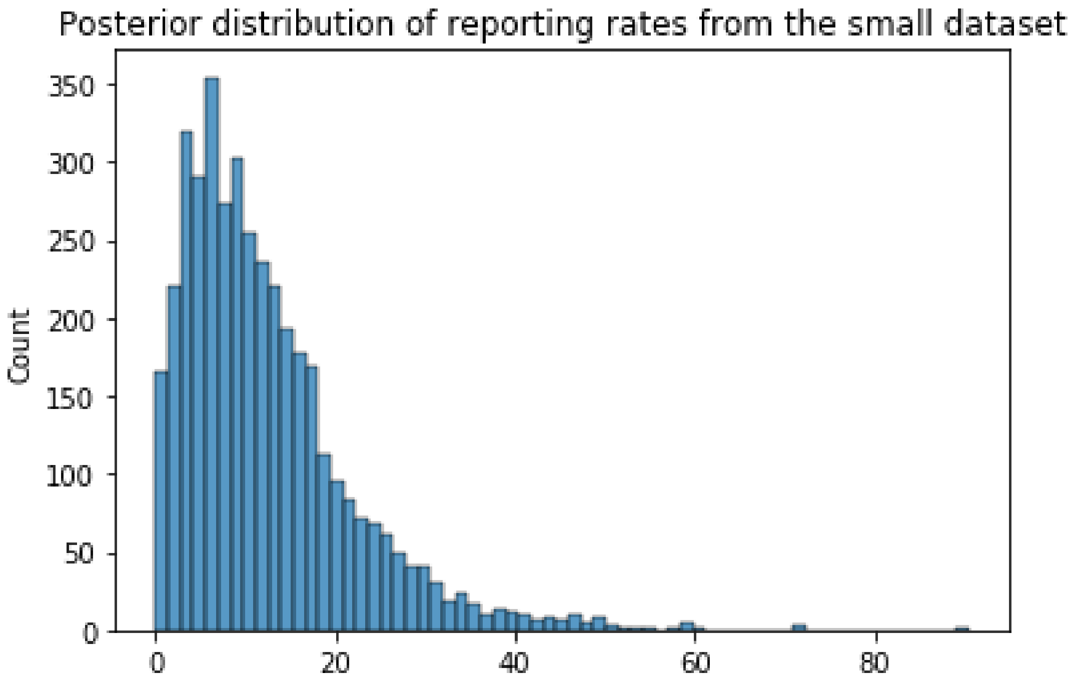


[30]:
```
sns.histplot(x=idata.posterior['reference_rate'].data.flatten())
plt.title('Posterior distribution of reporting rates from the complete dataset')
plt.show()
```

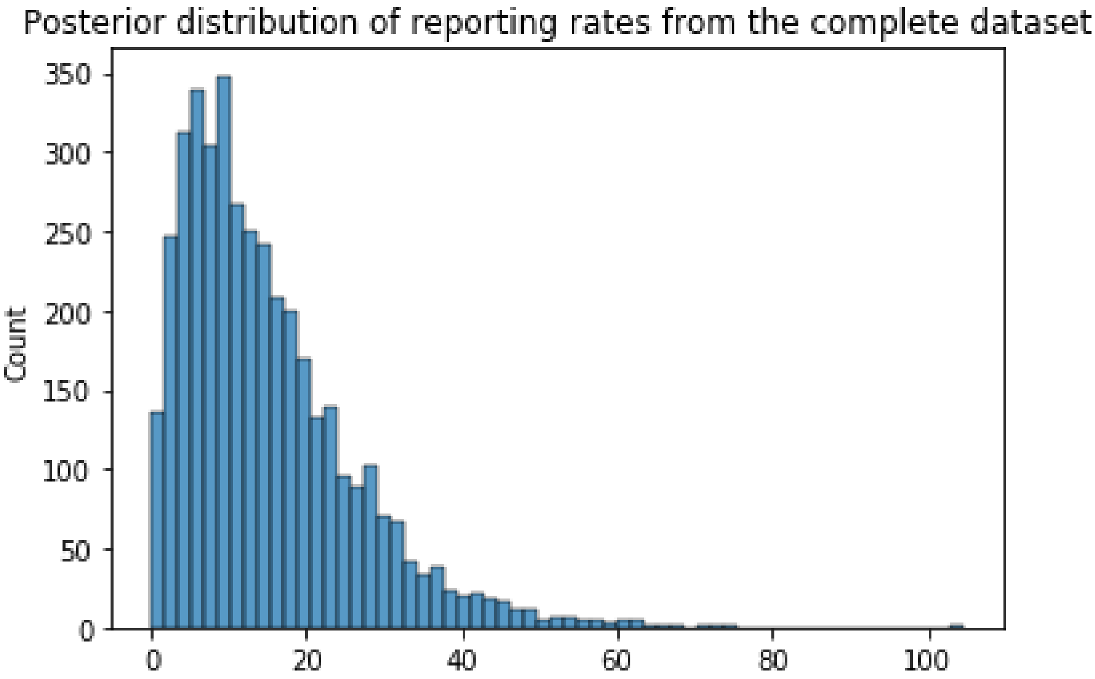
Posterior distribution of reporting rates from the complete dataset
Count
350
300
250
200
150
100
50
0
0
20
40
60
80
100

```
[ ]:
```

```
[ ]:
```
